# Revisiting the Definition of Acral Melanoma: Unraveling Histology and Location

**DOI:** 10.1101/2025.06.24.25330235

**Authors:** Emma Beagles, Sara Khattab, Olivia M. Burke, Arjun Mahajan, Danica Ciampa, Suzanne Xu, Christopher Thang, Cameron Moseley, Guihong Wan, Crystal Chang, Charles Lu, Nga Nguyen, Rebecca I. Hartman, Maryam M. Asgari, Mia DeSimone, Marc S. Hurlbert, Yevgeniy R. Semenov

## Abstract

Acral melanoma (AM) is a rare subtype of melanoma associated with poor prognosis. However, inconsistent definitions—based on either anatomic location (e.g., palms, soles, subungual areas) or histopathologic subtype (e.g., acral lentiginous melanoma)—complicate prognostication, hinder treatment decision-making, and pose challenges for both clinical and translational research. This multi-institutional retrospective cohort study aimed to determine whether anatomic location or histologic subtype better predicts outcomes in AM. Distal extremity primary melanomas (n = 469) were matched 1:2 with cutaneous melanomas (CMs; n = 938). Cox proportional hazards models evaluated recurrence-free survival (RFS), melanoma-specific survival (MSS), and overall survival (OS). Of the distal tumors, 280 (59.7%) were ALMs, including 33 (11.8%) on non-subungual dorsal sites. Compared to CMs, acral surface melanomas had worse outcomes, while dorsal melanomas had similar outcomes, independent of histology. ALMs were associated with worse survival than superficial spreading melanomas (SSMs). In interaction models, ALMs on dorsal surfaces and all histologic subtypes on acral surfaces had worse MSS than non-distal SSMs. A revised definition of AM may better inform clinical management and future research.

## Introduction

Acral melanoma (AM) is a rare subtype of melanoma with poor prognosis.^1^ Despite being one of the most common subtypes of melanoma in individuals with skin of color,^2^ AM remains understudied compared to other melanoma forms. A significant barrier to advancing the understanding of AM lies in the lack of standardized diagnostic criteria—specifically, whether AM should be defined by anatomic location (e.g., palms, soles, and subungual regions) or by histopathologic subtype (e.g., a lentiginous growth pattern).^3–6^ Indeed, a recent review highlighted the significant challenge posed by inconsistent definitions: only 38% of included studies reported the histopathological subtypes studied, 78% reported anatomical sites, 37% reported both, and 21% provided no specification of either.^7^ As a result, the current inconsistent classification system hinders accurate prognostication and treatment selection and serves as an obstacle to basic and clinical research.

In this study, we leverage a large multi-institutional melanoma registry to propose a refined approach for defining AMs according to patient outcomes. Specifically, we examine the survival among patients with distal extremity melanomas to understand the extent to which these outcomes are driven by histologic behavior or tumor location.

## Results

A total of 469 distal extremity melanomas were identified and matched to 938 CMs. Of the distal extremity melanomas, 362 (77.2%) were on acral locations and 107 (22.8%) were on dorsal locations. Notably, 280 (59.7%) of the distal extremity melanomas were histologically confirmed ALMs, of which 33 (11.8%) occurred on non-subungual dorsal locations. There were more White patients in CMs (932/938 [99.4%]) compared to acral (316/362 [87.3%]) and dorsal (96/107 [89.7%]) location melanomas (p<0.001). Melanomas on acral locations were diagnosed at a later stage (median [IQR], 2 [1-3]) compared to dorsal location melanomas (1 [1-2]) (p=0.002). Patients with acral location melanomas were more likely to have recurrence and had shorter survival compared to those with dorsal location melanomas and CMs, regardless of histologic subtype (Table I and eTable I).

**Table I.** Demographics and tumor-related information by location of primary melanoma.

|  | **Acral,<sup>a</sup> n=362** | **Dorsal,<sup>b</sup> n=107** | **CM, n=938** | **p-value** |
| --- | --- | --- | --- | --- |
| **Age, mean (SD)** | 62.2 (16.1) | 54.3 (18.9) | 59.5 (16.2) | 0.078 |
| **Male Sex, n (%)** | 154 (42.5) | 39 (36.5) | 405 (43.2) | 0.411 |
| **Race, n (%)** |  |  |  |  |
| American Indian or Alaska Native | 0 (0) | 1 (0.9) | 0 (0) | <0.001 |
| Asian | 4 (1.1) | 5 (4.7) | 2 (0.2) |  |
| Black | 18 (5.0) | 2 (1.9) | 0 (0) |  |
| Native Hawaiian or Pacific Islander | 2 (0.6) | 0 (0) | 0 (0) |  |
| Other<sup>c</sup> | 6 (1.6) | 0 (0) | 2 (0.2) |  |
| White | 316 (87.3) | 96 (89.7) | 932 (99.4) |  |
| Unknown | 16 (4.4) | 3 (2.8) | 2 (0.2) |  |
| **Ethnicity, n (%)** |  |  |  |  |
| Non-Hispanic | 283 (78.2) | 86 (90.4) | 882 (94.0) | <0.001 |
| Hispanic | 15 (4.1) | 2 (1.9) | 3 (0.3) |  |
| Unknown | 64 (17.7) | 19 (17.7) | 53 (5.7) |  |
| **CCI, median (IQR)** | 0 (0-1) | 0 (0-0) | 0 (0-1) | 0.007 |
| **Stage, median (IQR)** | 2 (1-3) | 1 (1-2) | 2 (1-2) | 0.002 |
| **Histology type, n (%)** |  |  |  |  |
| Acral lentiginous melanoma | 247 (68.2) | 32 (29.9) | 0 (0) | <0.001 |
| Lentigo maligna | 1 (0.3) | 0 (0) | 62 (6.6) |  |
| Lentigo maligna melanoma | 0 (0) | 2 (1.9) | 51 (5.4) |  |
| Malignant melanoma NOS | 72 (19.9) | 33 (30.8) | 241 (25.7) |  |
| Nodular melanoma | 26 (7.2) | 7 (6.6) | 180 (19.2) |  |
| Superficial spreading melanoma | 16 (4.4) | 33 (30.8) | 404 (43.1) |  |
| **Recurrence Type, n (%)** |  |  |  |  |
| Present | 140 (38.7) | 22 (20.6) | 272 (29.0) | <0.001 |
| *Distant* | 48 (13.3) | 5 (4.7) | 145 (15.5) |  |
| *Local* | 92 (25.4) | 17 (15.9) | 127 (13.5) |  |
| Never disease-free | 22 (6.1) | 2 (4.8) | 44 (4.7) |  |
| Absent | 200 (55.3) | 83 (77.6) | 622 (66.3) |  |
| **Melanoma-specific mortality, n (%)** | 91 (25.1) | 14 (13.1) | 159 (16.9) | 0.001 |
| **All-cause mortality, n (%)** | 159 (43.9) | 25 (23.4) | 334 (35.6) | <0.001 |
| **Survival time (months), median (IQR)** | 65 (28-118) | 91 (46-147) | 82 (32-142) | 0.005 |
| <sup>a</sup>Acral includes melanomas on palmar, plantar, and subungual locations |  |  |  |  |
| <sup>b</sup>Dorsal includes melanomas on non-subungual dorsal surfaces of the hands and feet |  |  |  |  |
| <sup>c</sup>Other was chosen for patients who manually described their race as “other” |  |  |  |  |
| CM=Cutaneous melanoma on non-distal extremity; SD=Standard deviation; IQR=Interquartile range; |  |  |  |  |
| CCI=Charlson Comorbidity Index; NOS=Not otherwise specified |  |  |  |  |

In univariate analyses, when examining histologic behavior, patients with both ALMs and NMs (regardless of location) had greater rates of recurrence and increased all-cause mortality than those with other histologic subtypes. ALMs (median [IQR], 64 [28-118]) had the shortest survival time followed by NMs (72 [27-123] months), melanoma NOS (78 [33-131]), SSMs (85 [28-150]), and Other (106 [62-170]) (p<0.001) (eTable II).

KM curves for RFS by location demonstrated that melanomas on acral locations performed worse than CMs, while dorsal location melanomas performed similar to CMs (Figure 2a). KM curves by histology showed increasingly worse RFS in the following order: Other (see above), SSM, Melanoma NOS, ALM, NM (Figure 2b) independent of location. These findings were consistent for MSS and OS (eFigures 1 and 2, respectively). KM curves for the interaction of location and histologic classification demonstrate that acral location melanomas generally performed worse for all outcomes compared to CMs independent of histologic subtype, with ALMs on dorsal locations also performing worse than CMs (eFigure 3a-c).

The complete multivariable models by location and histologic classification are shown in eTables III and IV, respectively. In analyses by location, acral location was independently associated with worse RFS (HR=1.31; CI=1.04-1.63; p=0.019), MSS (HR=2.12; CI=1.59-2.82; p<0.001), and OS (HR=1.46; CI=1.18-1.80; p<0.001) compared to CMs, while dorsal locations performed similar to CMs for all outcomes. For histologic analyses, ALMs had worse RFS (HR=1.46; CI=1.09-1.94; p=0.010), MSS (HR=1.88; CI=1.29-2.74; p=0.001), and OS (HR= 1.33; CI=1.02-1.73; p=0.032) compared to all SSMs. NMs also had worse RFS compared to all SSMs (HR=1.64; CI=1.24-2.18; p=0.001).

Analyses assessing the interaction between location and histology demonstrated that on acral surfaces, ALMs (HR=1.52; CI=1.12-2.07; p=0.007) and NMs (HR=2.61; CI=1.51-4.51; p=0.001) had worse RFS compared to SSMs on non-distal extremities. Moreover, all histologic subtypes of melanoma on acral surfaces exhibited worse MSS than SSMs on non-distal extremities. Notably, dorsal location ALMs had worse MSS compared to SSMs on non-distal extremities (HR=2.22; CI=1.03-4.81; p=0.042). Acral surface ALMs also had worse OS (HR=1.34; CI=1.01-1.77; p=0.039) compared to SSMs on non-distal extremities (Table II).

**Table II.**
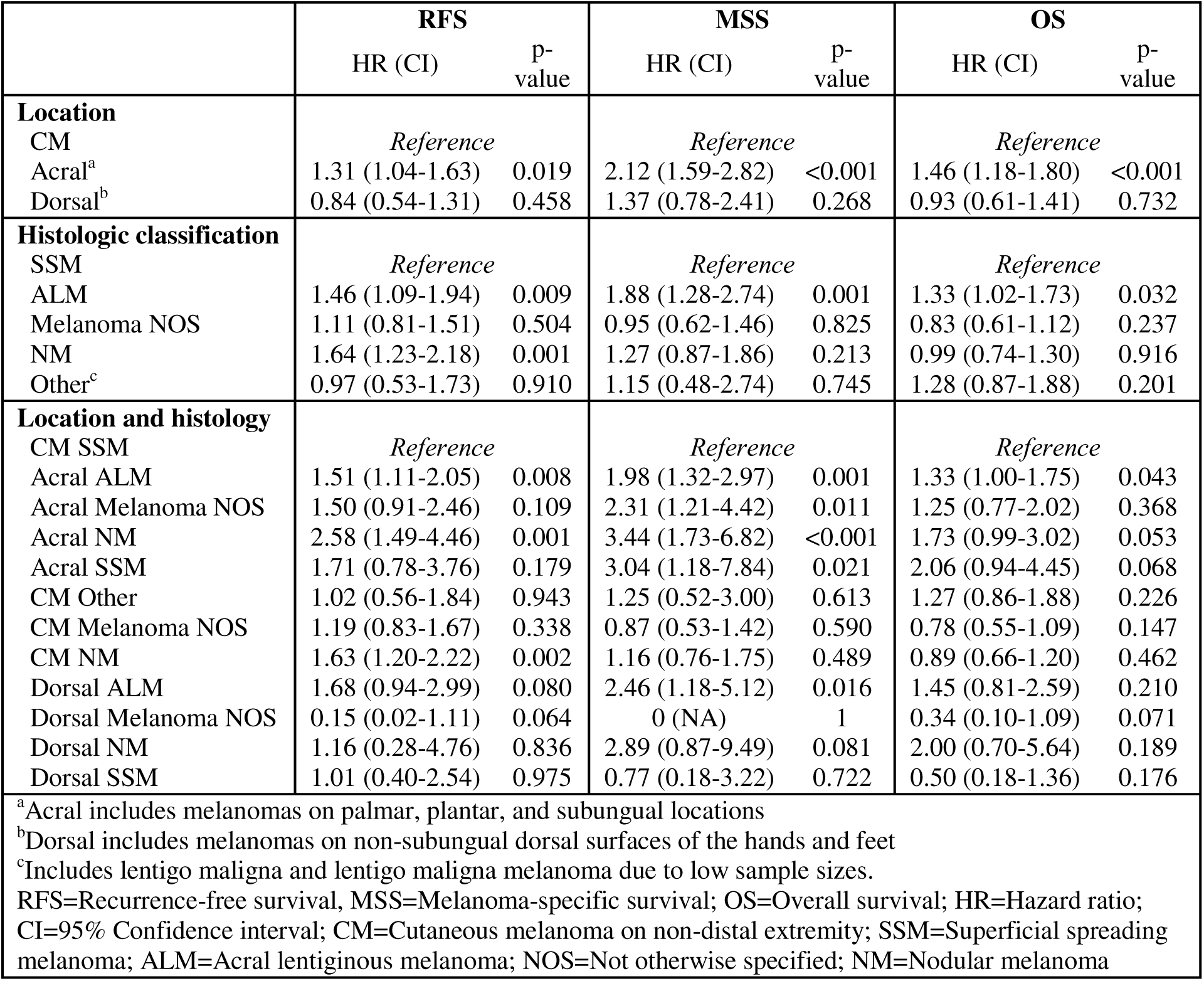
Multivariable Cox proportional hazard regressions for RFS, MSS, and OS by melanoma location, histologic classification, and their interaction adjusted for age, sex, race, CCI, stage, and diagnosis year.

## Discussion

This multi-institutional retrospective cohort study offers a novel and granular analysis of AM, leveraging a large dataset with detailed clinical annotations and long-term survival outcomes— features missing from publicly available cancer registries. Our findings underscore the urgent need to refine the current definition of AM, which remains inconsistently applied across clinical and research contexts.

Traditionally, AM has been defined by either anatomic location or histologic subtype. However, our data challenge the adequacy of this framework. We observed that 31.8% of melanomas on acral surfaces were not histologically classified as ALM, while 29.9% of melanomas on dorsal surfaces of the distal extremities were classified as ALM. Prior reports have similarly documented the presence of histologic ALM on non-acral surfaces.^8^ These findings directly dispute the prevailing diagnostic approach that requires ALMs to arise on acral surfaces,^9^ and suggest that biologically aggressive ALM may be defined by features beyond anatomic restriction.

Our study demonstrates that melanomas on acral surfaces—regardless of histologic subtype— consistently exhibited poorer survival outcomes compared to cutaneous melanomas on non-distal extremities. Conversely, melanomas on dorsal surfaces generally behaved more like cutaneous melanomas, with the exception of dorsal ALMs, which had similarly poor MSS as their acral counterparts. These findings suggest that location and histology each contribute prognostic value and that their overlap—particularly ALM on acral or dorsal surfaces—may identify a particularly high-risk subgroup.

Importantly, although our data and prior studies show that acral melanomas tend to be diagnosed at later stages,^1,10,11^ the inferior outcomes associated with acral surface melanomas persisted even after adjusting for stage. As a result, we conclude that other factors must be contributing to these poorer survival outcomes. Specifically, we suspect melanomas occurring on acral locations may be more aggressive due to differences in immune surveillance on these surfaces,^12,13^ as well as non-UV driven mechanisms of cancer development on the palms and soles (e.g., repetitive trauma).^14,15^

A limitation of our study is the potential for histologic misclassification by diagnosing pathologists—for example, labeling non-ALMs on acral surfaces or ALMs on dorsal surfaces. However, in these instances, the pathologist knowingly deviated from the most common histologic subtype for that location, suggesting heightened diagnostic confidence. Moreover, ALM remains the most common histopathologic subtype of acral melanoma followed by NM and SSM,^16,17,18^ supporting the plausibility of the observed distributions.

Our findings provide support for a refined framework for defining AM—one that is guided by survival outcomes rather than anatomic or histologic criteria alone. Notably, acral surface melanomas and dorsal surface ALMs may represent a distinct group warranting closer surveillance and tailored management strategies. Moving forward, molecular profiling of melanomas across acral and dorsal surfaces is needed to investigate the genomic, transcriptomic, and immunologic mechanisms that drive these divergent behaviors. While Wang et al. explored the genomic evolution of AM, their analyses did not stratify by histology or surface location.^19^ Stratifying future analyses by both histologic subtype and precise anatomic location will be critical, particularly given emerging data suggesting that AMs vary in immune infiltration and mutational signatures.

Ultimately, integrating biologic and anatomic insights will be essential to advance AM classification, guide future research, and improve patient outcomes. Our study lays the groundwork for this evolution in definition and encourages further investigations to validate and refine this approach through molecular phenotyping and prospective outcome correlations.

## Methods

Patients seen within the Mass General Brigham Healthcare System and Dana-Farber Cancer Institute with a primary melanoma from 1980-2020 meeting either of the following criteria were included in the study: 1) International Classification of Diseases for Oncology 3rd Edition (ICD-O-3) histologic behavior code for acral lentiginous melanoma (ALM) 87443 or C44.x independent of tumor location, or 2) melanomas on the hands, feet, or nail unit independent of ICD-O-3 histologic behavior. All cases were independently adjudicated via manual chart review to confirm the melanoma histologic subtype (e.g., superficial spreading melanoma (SSM), nodular melanoma (NM), ALM, melanoma not otherwise specified (NOS)) and location on the distal extremities (acral if on palmar, plantar, or subungual areas vs dorsal locations of hands and feet). To establish acral vs dorsal location in ambiguous cases, chart review included review of photographs taken and surgical operative reports to confirm surface classification with certainty. We excluded 7 melanomas due to unknown stage and 3 ALMs by ICD-O-3 histologic behavior due to non-distal extremity location of the primary melanoma, thought to be consistent with cutaneous metastases from regressed primaries or misclassification. Additionally, all histologically designated ALM on dorsal surface and SSM on acral surface pathologic specimens were reviewed under the microscope by a board-certified dermatopathologist to confirm the diagnosing pathologist’s designations. Distal extremity melanomas were 1:2 propensity-score matched with primary cutaneous melanomas on non-distal extremities (CM) based on stage, age, sex, and diagnosis year to provide a comparator cohort for benchmarking survival outcomes (Figure 1). Manual chart review was performed to extract mortality status, cause of death, recurrence status and type, date of last contact, histopathologic features, and last known cancer status. All manual chart review was conducted by two independent reviewers with a third reviewer reconciling any differences. Lentigo maligna and lentigo maligna melanoma were grouped into “Other” for histologic analyses due to their low sample sizes.

**Figure 1.**
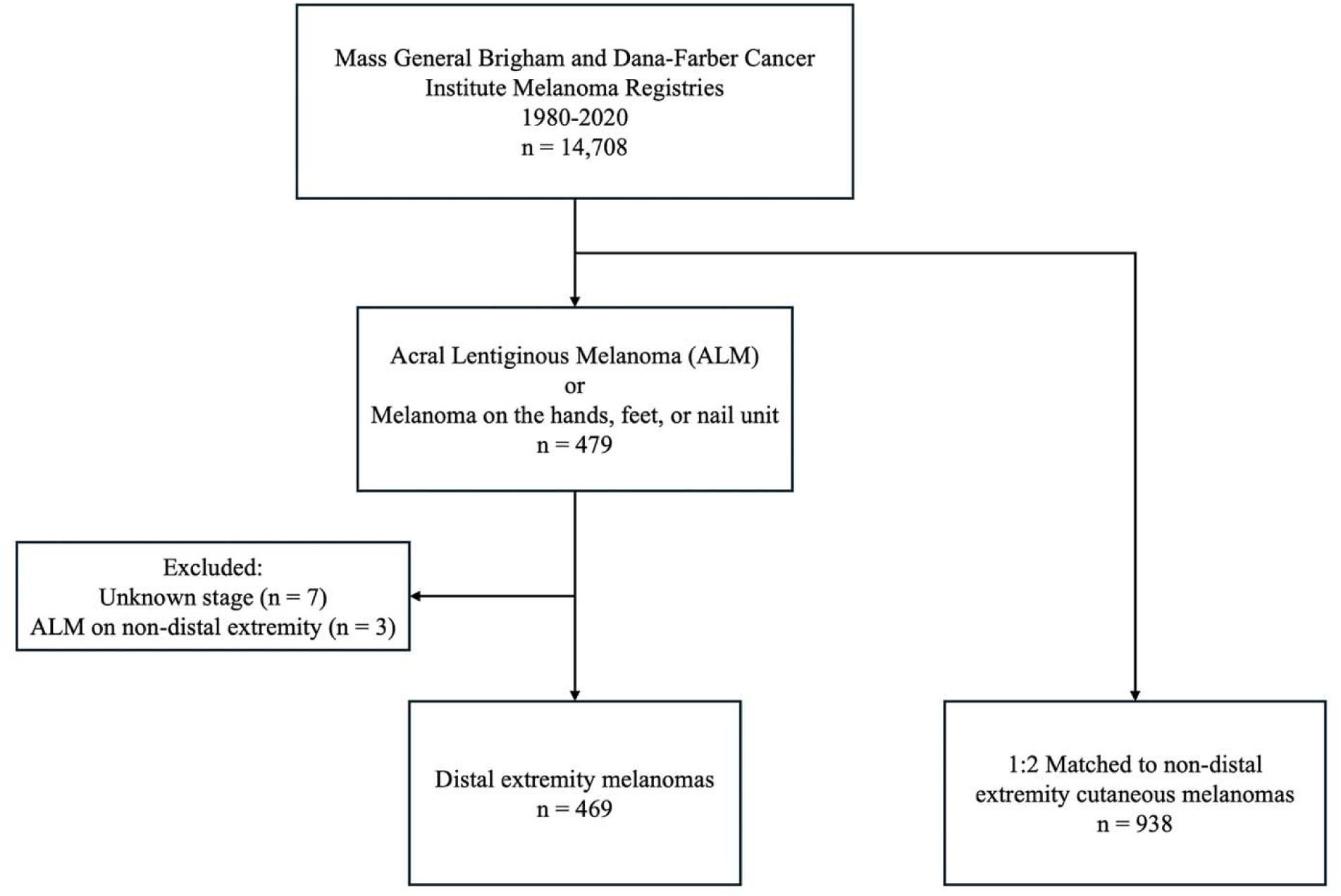
Study flow chart of cases and controls for distal extremity melanomas and cutaneous melanomas on non-distal extremities.

**Figure 2.**
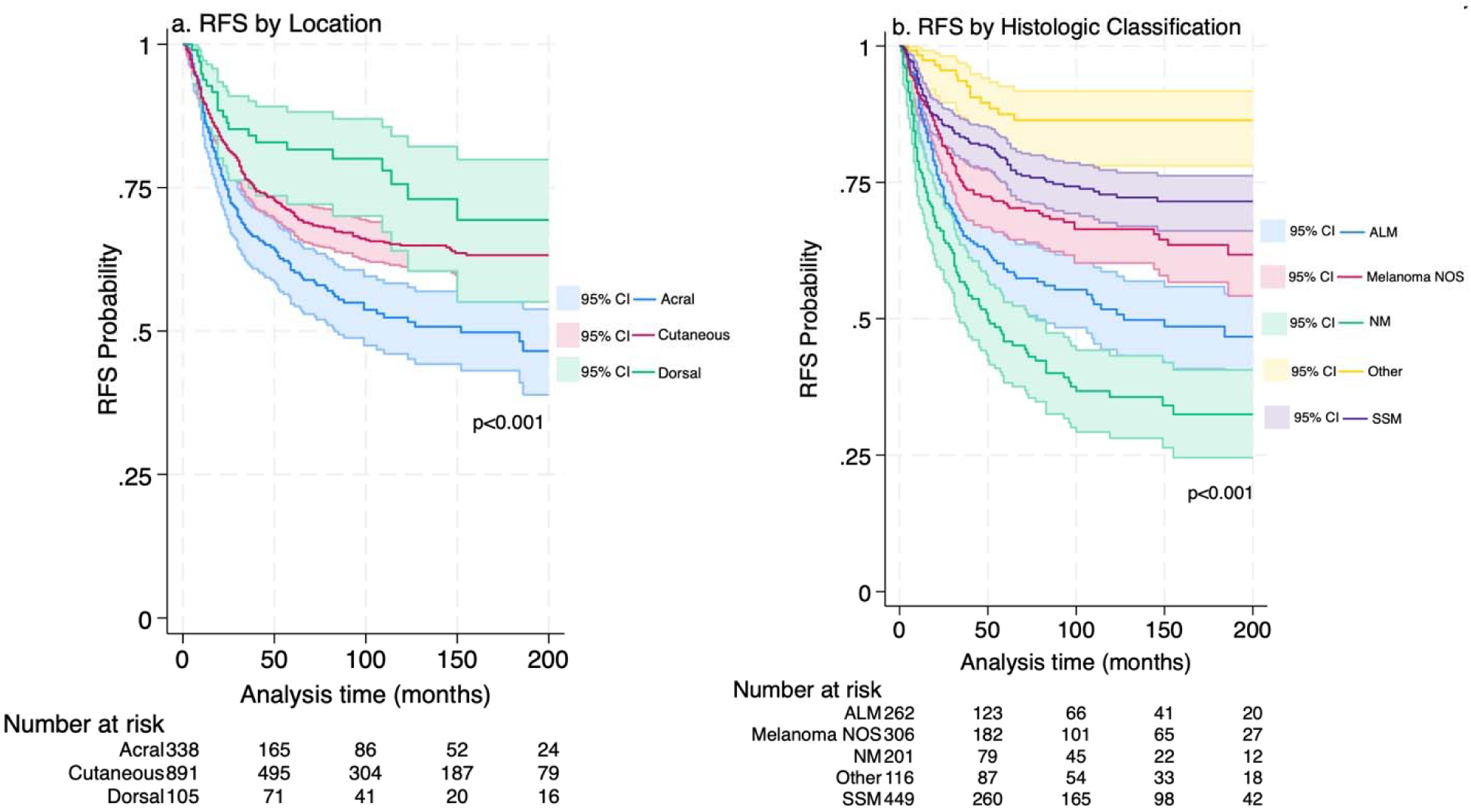
Kaplan-Meier curves for recurrence-free survival (RFS) by (a) location of melanoma and (b) histologic classification of melanoma. Acral location includes melanomas on palmar, plantar, and subungual surfaces. Dorsal location includes melanomas on non-subungual dorsal surfaces of the hands and feet. Cutaneous location refers to cutaneous melanomas on non-distal extremities. “Other” histology includes lentigo maligna and lentigo maligna melanoma due to low sample sizes. Acronyms: ALM=Acral lentiginous melanoma; NOS=Not otherwise specified; NM=Nodular melanoma; SSM=Superficial spreading melanoma.

Patient characteristics were compared in univariate analyses using Pearson’s ^2^ test for categorical variables, analysis of variance for continuous variables, and Kruskal-Wallis test for ordinal data. Kaplan-Meier (KM) curves were utilized to investigate differences in recurrence-free survival (RFS), melanoma-specific survival (MSS), and overall survival (OS) between patients with different melanoma locations and histologic behavior. Multivariable Cox proportional hazards regression models were used to examine the impact of melanoma location, histologic classification, and the interaction between location and histologic classification on RFS, MSS, and OS, adjusting for age, sex, race, Charlson Comorbidity Index (CCI), stage, and diagnosis year. SSM was used as the histologic subtype reference group, as it is the most common type of melanoma in our cohort. Due to low sample size of patients who were not White, race was categorized into “White,” “Non-White,” and “Unknown” in multivariable regressions. A two-sided P-value<0.05 was considered statistically significant. Statistical analyses were conducted using Stata 18.0 (StataCorp).

## Ethics StatementApproval

Reviewed and approved by Mass General Brigham Institutional Review Board (Protocol #2020P002113)

## Supporting information

STROBE Checklist

## Data availability statement

The data that support the findings of this study are available from the corresponding author upon reasonable request.

