## Supplement for "Revisiting the Definition of Acral Melanoma: Unraveling Histology and Location"

**Supplemental Material for “Revisiting the Definition of Acral Melanoma: Unraveling Histology and Location”**

**eTables: 4**

eTable I. Anatomic site of primary melanoma by location.

eTable II. Demographics and tumor-related information by histologic subtype of primary melanoma.

eTable III. Multivariable Cox proportional hazard regressions for recurrence-free survival (RFS), melanoma-specific survival (MSS), and overall survival (OS) for location of melanoma.

eTable IV. Multivariable Cox proportional hazard regressions for recurrence-free survival (RFS), melanoma-specific survival (MSS), and overall survival (OS) for histologic classification of melanoma.

eFigure 3c. Kaplan-Meier curves by the interaction between location and histologic classification of melanoma for overall survival (OS). Acral location includes melanomas on palmar, plantar, and subungual surfaces. Dorsal location includes melanomas on non-subungual dorsal surfaces of the hands and feet. Cutaneous location refers to cutaneous melanomas on non-distal extremities. “Other” histology includes lentigo maligna and lentigo maligna melanoma due to low sample sizes. Acronyms: ALM=Acral lentiginous melanoma; NOS=Not otherwise specified; NM=Nodular melanoma; SSM=Superficial spreading melanoma.

**eTable I**. Anatomic site of primary melanoma by location.

|  | **Acral,^a^ n=362** | **Dorsal,^b^ n=107** | **CM, N=938** | **p-value** |
| --- | --- | --- | --- | --- |
| **Site, n (%)** |  |  |  | <0.001 |
| Non-distal extremity | 0 (0) | 0 (0) | 938 (100.0) |  |
| Foot | 236 (65.2) | 89 (83.2) | 0 (0) |  |
| Hand | 23 (6.5) | 18 (16.8) | 0 (0) |  |
| Subungual foot | 50 (13.8) | 0 (0) | 0 (0) |  |
| Subungual hand | 53 (14.6) | 0 (0) | 0 (0) |  |
| ^a^Acral includes melanomas on palmar, plantar, and subungual locations  ^b^Dorsal includes melanomas on non-subungual dorsal surfaces of the hands and feet  CM=Cutaneous melanoma on non-distal extremity | | | | |

**eTable II.** Demographics and tumor-related information by histologic subtype of primary melanoma.

|  | **ALM,**  **N=279** | **Melanoma NOS,**  **N=346** | **NM,**  **N=213** | **SSM, N=453** | **Other,^a^**  **N=116** | **p-value** |
| --- | --- | --- | --- | --- | --- | --- |
| **Age at diagnosis, mean (SD)** | 62.6 (15.6) | 58.7 (18.0) | 61.4 (16.2) | 57.8 (15.6) | 61.6 (17.2) | 0.039 |
| **Male Sex, n (%)** | 120 (43.0) | 145 (41.9) | 112 (52.6) | 167 (36.9) | 54.0 (46.6) | 0.004 |
| **Race, n (%)** |  |  |  |  |  | <0.001 |
| American Indian or Alaska Native | 0 (0) | 0 (0) | 0 (0) | 1 (0.2) | 0 (0) |  |
| Asian | 5 (1.8) | 3 (0.9) | 0 (0) | 3 (0.7) | 0 (0) |  |
| Black | 16 (5.7) | 3 (0.9) | 0 (0) | 1 (0.2) | 0 (0) |  |
| Native Hawaiian or Pacific Islander | 1 (0.4) | 1 (0.3) | 0 (0) | 0 (0) | 0 (0) |  |
| Other^b^ | 2 (0.7) | 3 (0.9) | 2 (0.9) | 1 (0.2) | 0 (0) |  |
| White | 247 (88.5) | 326 (94.2) | 209 (98.1) | 446 (98.5) | 116 (100) |  |
| Unknown | 8 (2.9) | 10 (2.9) | 2 (0.9) | 1 (0.2) | 0 (0) |  |
| **Ethnicity, n (%)** |  |  |  |  |  | <0.001 |
| Non-Hispanic | 225 (80.6) | 293 (84.7) | 197 (92.5) | 431 (95.1) | 105 (90.5) |  |
| Hispanic | 13 (4.7) | 3 (0.9) | 14 (6.6) | 2 (0.4) | 11 (9.5) |  |
| Unknown | 41 (14.7) | 50 (14.5) | 2 (0.9) | 20 (4.4) | 0 (0) |  |
| **CCI** |  |  |  |  |  |  |
| Median (IQR) | 0 (0-1) | 0 (0-1) | 0 (0-1) | 0 (0-1) | 0 (0-1) | 0.297 |
| **Stage, median (IQR)** | 2 (1-3) | 2 (0-3) | 2 (2-3) | 1 (1-2) | 0 (0-1) | <0.001 |
| **Location, n (%)** |  |  |  |  |  | <0.001 |
| Acral^c^ | 247 (88.5) | 72 (20.8) | 26 (12.2) | 16 (3.5) | 1 (0.9) |  |
| Dorsal^d^ | 32 (11.5) | 33 (9.5) | 7 (3.3) | 33 (7.3) | 2 (1.7) |  |
| CM | 0 (0) | 241 (69.7) | 180 (84.5) | 404 (89.2) | 113 (97.4) |  |
| **Site, n (%)** |  |  |  |  |  | <0.001 |
| Non-distal extremity | 0 (0) | 241 (69.7) | 180 (84.5) | 404 (89.2) | 113 (97.4) |  |
| Foot | 191 (68.5) | 70 (20.2) | 26 (12.2) | 36 (7.9) | 2 (1.7) |  |
| Hand | 18 (6.5) | 9 (2.6) | 2 (0.9) | 11 (2.4) | 1 (0.9) |  |
| Subungual hand | 37 (13.3) | 15 (4.3) | 1 (0.5) | 0 (0) | 0 (0) |  |
| Subungual foot | 33 (11.8) | 11 (3.2) | 4 (1.9) | 2 (0.4) | 0 (0) |  |
| **Recurrence Type, n (%)** |  |  |  |  |  | <0.001 |
| Present | 109 (39.0) | 96 (27.7) | 115 (54.0) | 100 (22.1) | 14 (12.0) |  |
| *Distant* | *35 (12.5)* | *52 (15.0)* | *57 (26.8)* | *47 (10.4)* | *7 (6.0)* |  |
| *Local* | *74 (26.5)* | *44 (12.7)* | *58 (27.2)* | *53 (11.7)* | *7 (6.0)* |  |
| Never disease-free | 16 (5.7) | 37 (10.7) | 11 (5.2) | 4 (0.9) | 0 (0) |  |
| Absent | 154 (55.2) | 213 (61.6) | 87 (40.8) | 349 (77.0) | 102 (87.9) |  |
| **Melanoma-specific mortality, n (%)** | 71 (25.5) | 71 (20.5) | 65 (30.5) | 51 (11.3) | 6 (5.2) | <0.001 |
| **All-cause mortality, n (%)** | 128 (45.9) | 114 (32.9) | 106 (49.8) | 130 (28.7) | 40 (34.5) | <0.001 |
| **Survival time (months), median (IQR)** | 65 (28-119) | 78 (33-131) | 72 (27-123) | 85 (27-150) | 106 (62-170) | <0.001 |
| ^a^Includes both lentigo maligna and lentigo maligna melanoma due to low sample sizes.  ^b^Other was chosen for patients who manually described their race as “other”  ^c^Acral includes melanomas on palmar, plantar, and subungual locations  ^d^Dorsal includes melanomas on non-subungual dorsal surfaces of the hands and feet  ALM=Acral lentiginous melanoma; NOS=Not otherwise specified; NM=Nodular melanoma; SSM=Superficial spreading melanoma; SD=Standard deviation; CCI=Charlson Comorbidity Index; IQR=Interquartile range; CM=Cutaneous melanoma on non-distal extremity | | | | | | |

**eTable III.** Multivariable Cox proportional hazard regressions for recurrence-free survival (RFS), melanoma-specific survival (MSS), and overall survival (OS) for location of melanoma.

|  | **RFS** | | **MSS** | | **OS** | |
| --- | --- | --- | --- | --- | --- | --- |
|  | **HR (CI)** | **p-value** | **HR (CI)** | **p-value** | **HR (CI)** | **p-value** |
| **Age at diagnosis** | 1.02 (1.01-1.02) | <0.001 | 1.02 (1.01-1.02) | <0.001 | 1.05 (1.04-1.05) | <0.001 |
| **Sex** |  |  |  |  |  |  |
| Male | *Reference* | *Reference* | *Reference* | *Reference* | *Reference* | *Reference* |
| Female | 0.76 (0.62-0.92) | 0.006 | 0.86 (0.66-1.11) | 0.249 | 0.82 (0.68-0.98) | 0.034 |
| **Race** |  |  |  |  |  |  |
| White | *Reference* | *Reference* | *Reference* | *Reference* | *Reference* | *Reference* |
| Non-White^a^ | 1.26 (0.72-2.19) | 0.406 | 0.96 (0.44-2.08) | 0.927 | 0.99 (0.58-1.69) | 0.980 |
| Unknown | 0.87 (0.35-2.12) | 0.763 | 0.80 (0.19-3.28) | 0.753 | 0.52 (0.16-1.62) | 0.259 |
| **CCI** | 1.02 (0.94-1.09) | 0.593 | 1.18 (1.08-1.27) | <0.001 | 1.15 (1.08-1.21) | <0.001 |
| **Stage Category** |  |  |  |  |  |  |
| 0 | *Reference* | *Reference* | 0 (NA) | NA | *Reference* | *Reference* |
| 1 | 2.87 (1.47-5.59) | 0.002 | *Reference^d^* | *Reference^d^* | 1.85 (1.20-2.81) | 0.005 |
| 2 | 13.5 (7.12-25.7) | <0.001 | 5.98 (3.80-9.40) | <0.001 | 4.02 (2.64-6.11) | <0.001 |
| 3 | 17.1 (8.99-32.5) | <0.001 | 9.97 (6.36-15.6) | <0.001 | 5.16 (3.37-7.88) | <0.001 |
| 4 | 14.1 (6.20-31.8) | <0.001 | 26.5 (15.8-44.4) | <0.001 | 10.7 (6.50-17.4) | <0.001 |
| **Diagnosis year range** |  |  |  |  |  |  |
| Prior to 2000 | *Reference* | *Reference* | *Reference* | *Reference* | *Reference* | *Reference* |
| 2000-2005 | 0.47 (0.26-0.82) | 0.009 | 1.35 (0.59-3.07) | 0.474 | 1.07 (0.63-1.78) | 0.802 |
| 2006-2010 | 0.57 (0.33-0.98) | 0.043 | 1.33 (0.59-2.99) | 0.488 | 1.09 (0.65-1.81) | 0.738 |
| 2011-2015 | 0.71 (0.41-1.21) | 0.212 | 1.28 (0.56-2.89) | 0.557 | 0.89 (0.52-1.49) | 0.663 |
| 2016-2020 | 0.75 (0.43-1.29) | 0.303 | 0.89 (0.38-2.07) | 0.789 | 0.68 (0.38-1.17) | 0.166 |
| **Location** |  |  |  |  |  |  |
| CM | *Reference* | *Reference* | *Reference* | *Reference* | *Reference* | *Reference* |
| Acral^b^ | 1.31 (1.04-1.63) | 0.019 | 2.12 (1.59-2.82) | <0.001 | 1.46 (1.18-1.80) | <0.001 |
| Dorsal^c^ | 0.84 (0.54-1.31) | 0.458 | 1.37 (0.78-2.41) | 0.268 | 0.93 (0.61-1.41) | 0.732 |
| ^a^Includes: American Indian or Alaska Native, Asian, Black, Native Hawaiian or Pacific Islander, and Other due to low sample sizes.  ^b^Acral includes melanomas on palmar, plantar, and subungual locations  ^c^Dorsal includes melanomas on non-subungual dorsal surfaces of the hands and feet  ^d^Stage 1 was used as the reference group for MSS due to the limited number of patients who initially had stage 0 disease and experienced melanoma-related mortality.  RFS=Recurrence-free survival, MSS=Melanoma-specific survival; OS=Overall Survival; HR=Hazard Ratio; CI=95% Confidence Interval; CCI=Charlson Comorbidity Index; CM=Cutaneous melanoma on non-distal extremity | | | | | | |

**eTable IV.** Multivariable Cox proportional hazard regressions for recurrence-free survival (RFS), melanoma-specific survival (MSS), and overall survival (OS) for histologic classification of melanoma.

|  | **RFS** | | **MSS** | | **OS** | |
| --- | --- | --- | --- | --- | --- | --- |
|  | **HR (CI)** | **p-value** | **HR (CI)** | **p-value** | **HR (CI)** | **p-value** |
| **Age at diagnosis** | 1.02 (1.01-1.03) | <0.001 | 1.02 (1.01-1.02) | <0.001 | 1.05 (1.04-1.05) | <0.001 |
| **Sex** |  |  |  |  |  |  |
| Male | *Reference* | *Reference* | *Reference* | *Reference* | *Reference* | *Reference* |
| Female | 0.77 (0.63-0.93) | 0.009 | 0.90 (0.69-1.16) | 0.434 | 0.85 (0.70-1.02) | 0.084 |
| **Race** |  |  |  |  |  |  |
| White | *Reference* | *Reference* | *Reference* | *Reference* | *Reference* | *Reference* |
| Non-White^a^ | 1.31 (0.75-2.27) | 0.345 | 1.02 (0.46-2.2) | 0.967 | 1.03 (0.59-1.75) | 0.922 |
| Unknown | 0.88 (0.35-2.13) | 0.772 | 0.94 (0.22-3.88) | 0.932 | 0.59 (0.18-1.85) | 0.362 |
| **CCI** | 1.01 (0.94-1.08) | 0.749 | 1.17 (1.07-1.26) | <0.001 | 1.15 (1.08-1.21) | <0.001 |
| **Stage Category** |  |  |  |  |  |  |
| 0 | *Reference* | *Reference* | 0 (NA) | NA | *Reference* | *Reference* |
| 1 | 2.78 (1.39-5.55) | 0.004 | *Reference^c^* | *Reference^c^* | 1.80 (1.14-2.81) | 0.011 |
| 2 | 11.6 (5.92-22.6) | <0.001 | 5.73 (3.58-9.19) | <0.001 | 4.16 (2.65-6.53) | <0.001 |
| 3 | 15.1 (7.68-29.6) | <0.001 | 9.61 (6.02-15.4) | <0.001 | 5.39 (3.40-8.53) | <0.001 |
| 4 | 13.4 (5.8-31.1) | <0.001 | 27.0 (15.0-48.6) | <0.001 | 12.3 (7.26-20.7) | <0.001 |
| **Diagnosis year range** |  |  |  |  |  |  |
| Prior to 2000 | *Reference* | *Reference* | *Reference* | *Reference* | *Reference* | *Reference* |
| 2000-2005 | 0.44 (0.25-0.76) | 0.004 | 1.08 (0.47-2.45) | 0.858 | 0.92 (0.55-1.53) | 0.750 |
| 2006-2010 | 0.55 (0.32-0.93) | 0.027 | 1.10 (0.48-2.47) | 0.823 | 0.97 (0.58-1.61) | 0.919 |
| 2011-2015 | 0.70 (0.41-1.18) | 0.181 | 1.05 (0.46-2.38) | 0.900 | 0.82 (0.48-1.36) | 0.446 |
| 2016-2020 | 0.78 (0.45-1.35) | 0.383 | 0.78 (0.33-1.82) | 0.571 | 0.63 (0.36-1.09) | 0.103 |
| **Histologic subtype** |  |  |  |  |  |  |
| SSM | *Reference* | *Reference* | *Reference* | *Reference* | *Reference* | *Reference* |
| ALM | 1.46 (1.09-1.94) | 0.009 | 1.88 (1.28-2.74) | 0.001 | 1.33 (1.02-1.73) | 0.032 |
| Melanoma NOS | 1.11 (0.81-1.51) | 0.504 | 0.95 (0.62-1.46) | 0.825 | 0.83 (0.61-1.12) | 0.237 |
| NM | 1.64 (1.23-2.18) | 0.001 | 1.27 (0.87-1.86) | 0.213 | 0.99 (0.74-1.30) | 0.916 |
| Other^b^ | 0.97 (0.53-1.73) | 0.910 | 1.15 (0.48-2.74) | 0.745 | 1.28 (0.87-1.88) | 0.201 |
| ^a^Includes: American Indian or Alaska Native, Asian, Black, Native Hawaiian or Pacific Islander, and Other due to low sample sizes.  ^b^Includes both lentigo maligna and lentigo maligna melanoma due to low sample sizes.  ^c^Stage 1 was used as the reference group for MSS due to the limited number of patients who initially had stage 0 disease and experienced melanoma-related mortality.  RFS=Recurrence-free survival, MSS=Melanoma-specific survival; OS=Overall survival; HR=Hazard ratio; CI=95% Confidence interval; CCI=Charlson Comorbidity Index; SSM=Superficial spreading melanoma; ALM=Acral lentiginous melanoma; NOS=Not otherwise specified; NM=Nodular melanoma | | | | | | |

**eFigure 1**. Kaplan-Meier curves for melanoma-specific survival (MSS) by (a) location of melanoma and (b) histologic classification of melanoma. Acral location includes melanomas on palmar, plantar, and subungual surfaces. Dorsal location includes melanomas on non-subungual dorsal surfaces of the hands and feet. Cutaneous location refers to cutaneous melanomas on non-distal extremities. “Other” histology includes lentigo maligna and lentigo maligna melanoma due to low sample sizes. Acronyms: ALM=Acral lentiginous melanoma; NOS=Not otherwise specified; NM=Nodular melanoma; SSM=Superficial spreading melanoma.
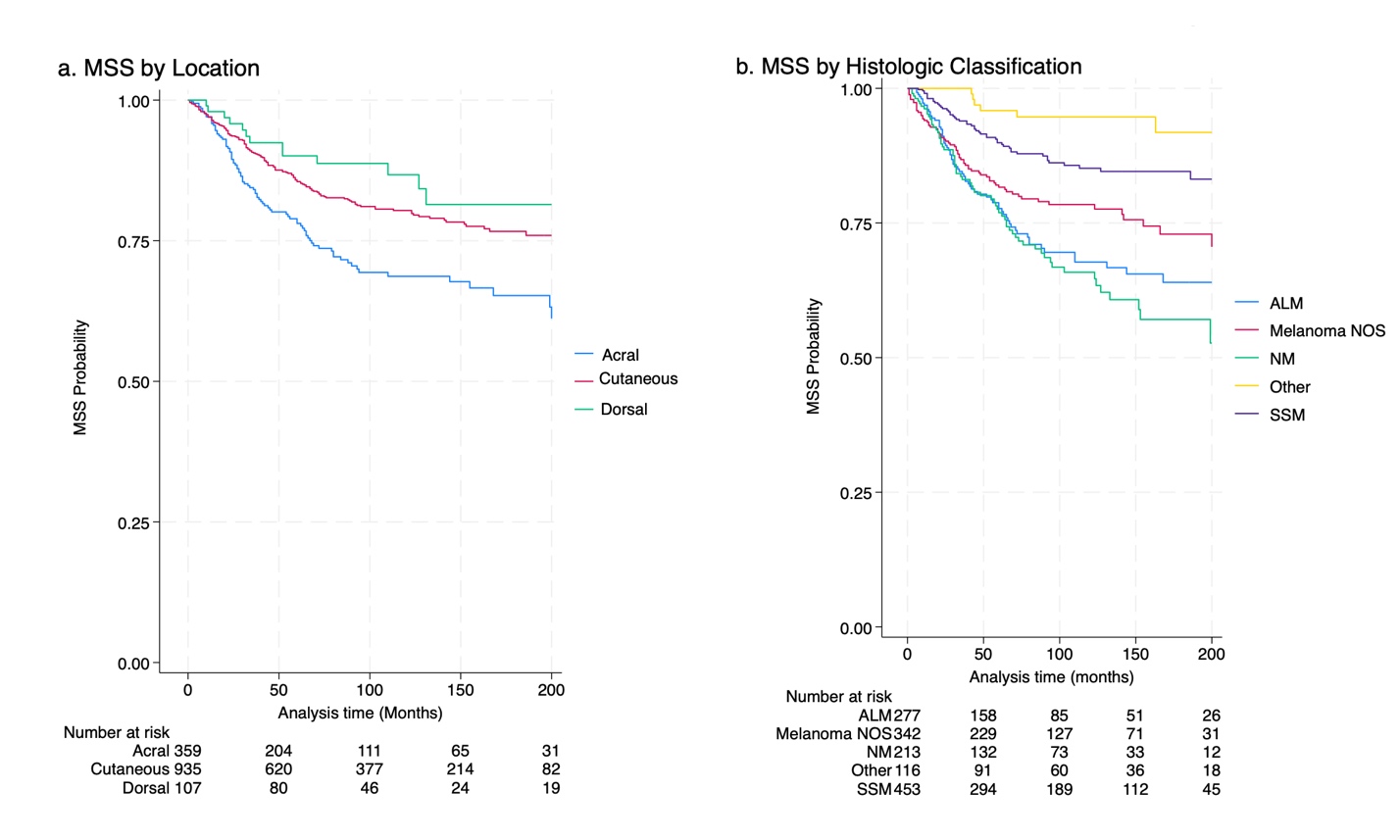

**eFigure 2**. Kaplan-Meier curves for overall survival (OS) by (a) location of melanoma and (b) histologic classification of melanoma. Acral location includes melanomas on palmar, plantar, and subungual surfaces. Dorsal location includes melanomas on non-subungual dorsal surfaces of the hands and feet. Cutaneous location refers to cutaneous melanomas on non-distal extremities. “Other” histology includes lentigo maligna and lentigo maligna melanoma due to low sample sizes. Acronyms: ALM=Acral lentiginous melanoma; NOS=Not otherwise specified; NM=Nodular melanoma; SSM=Superficial spreading melanoma.
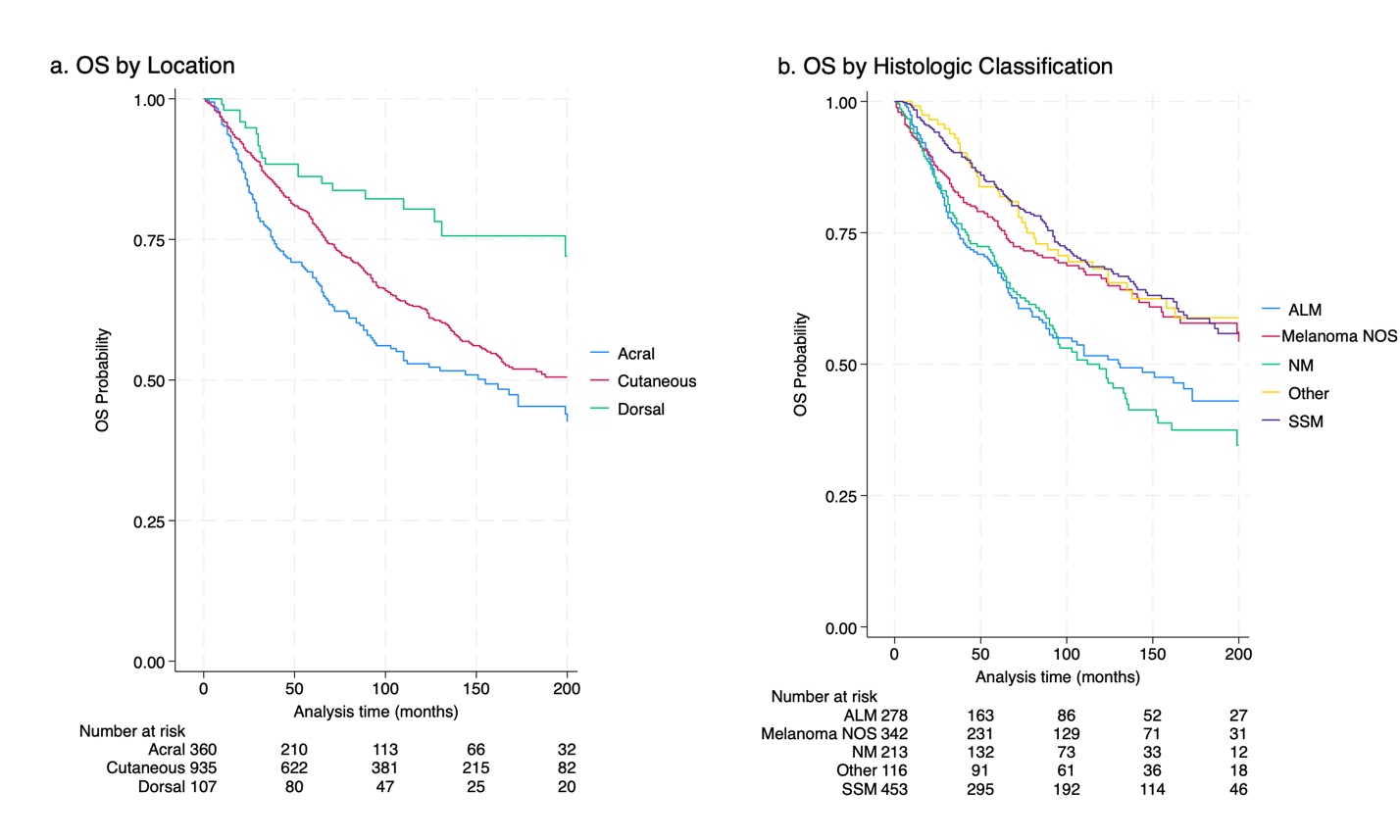

**eFigure 3a**. Kaplan-Meier curves by the interaction between location and histologic classification of melanoma for recurrence-free survival (RFS). Acral location includes melanomas on palmar, plantar, and subungual surfaces. Dorsal location includes melanomas on non-subungual dorsal surfaces of the hands and feet. Cutaneous location refers to cutaneous melanomas on non-distal extremities. “Other” histology includes lentigo maligna and lentigo maligna melanoma due to low sample sizes. Acronyms: ALM=Acral lentiginous melanoma; NOS=Not otherwise specified; NM=Nodular melanoma; SSM=Superficial spreading melanoma.

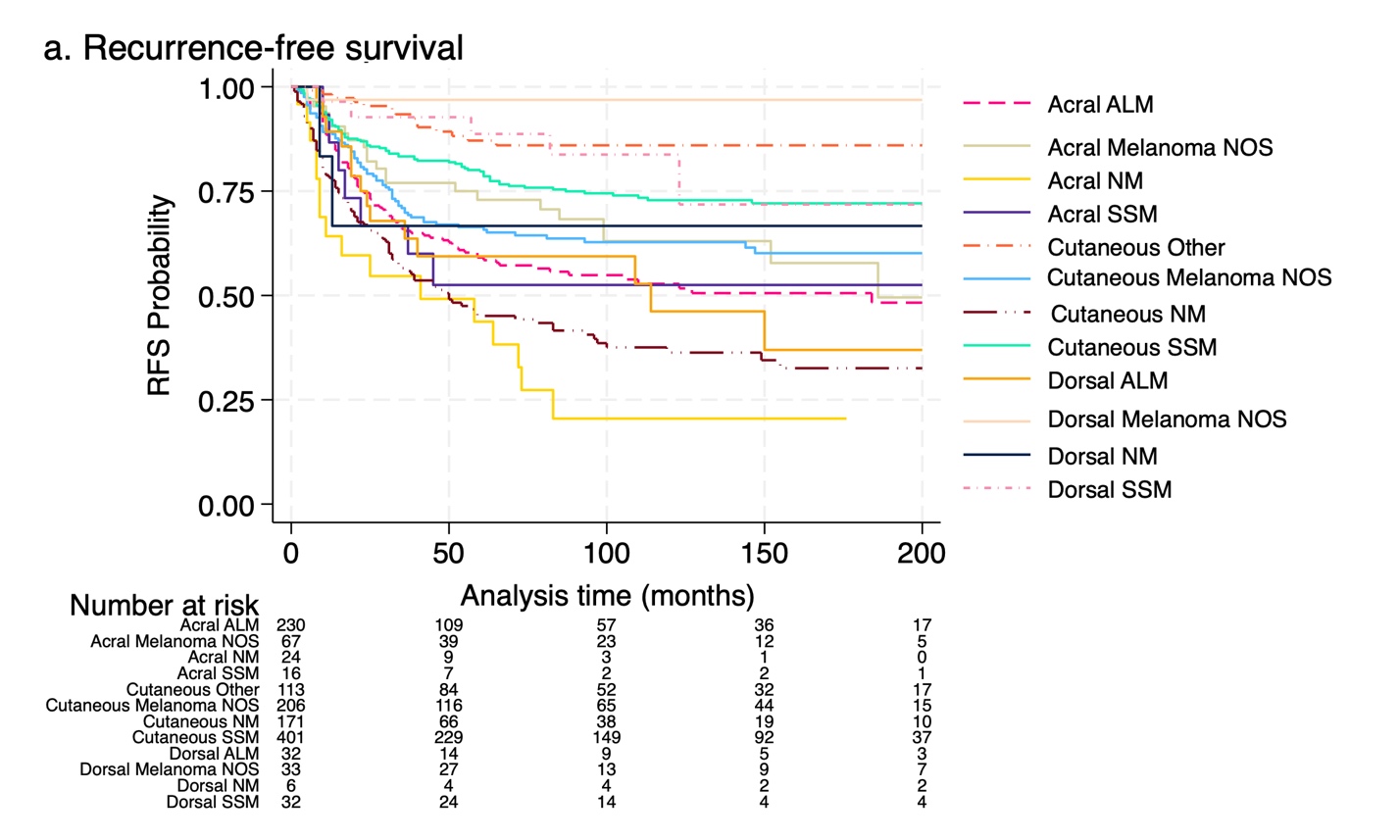

**eFigure 3b**. Kaplan-Meier curves by the interaction between location and histologic classification of melanoma for melanoma-specific survival (MSS). Acral location includes melanomas on palmar, plantar, and subungual surfaces. Dorsal location includes melanomas on non-subungual dorsal surfaces of the hands and feet. Cutaneous location refers to cutaneous melanomas on non-distal extremities. “Other” histology includes lentigo maligna and lentigo maligna melanoma due to low sample sizes. Acronyms: ALM=Acral lentiginous melanoma; NOS=Not otherwise specified; NM=Nodular melanoma; SSM=Superficial spreading melanoma.

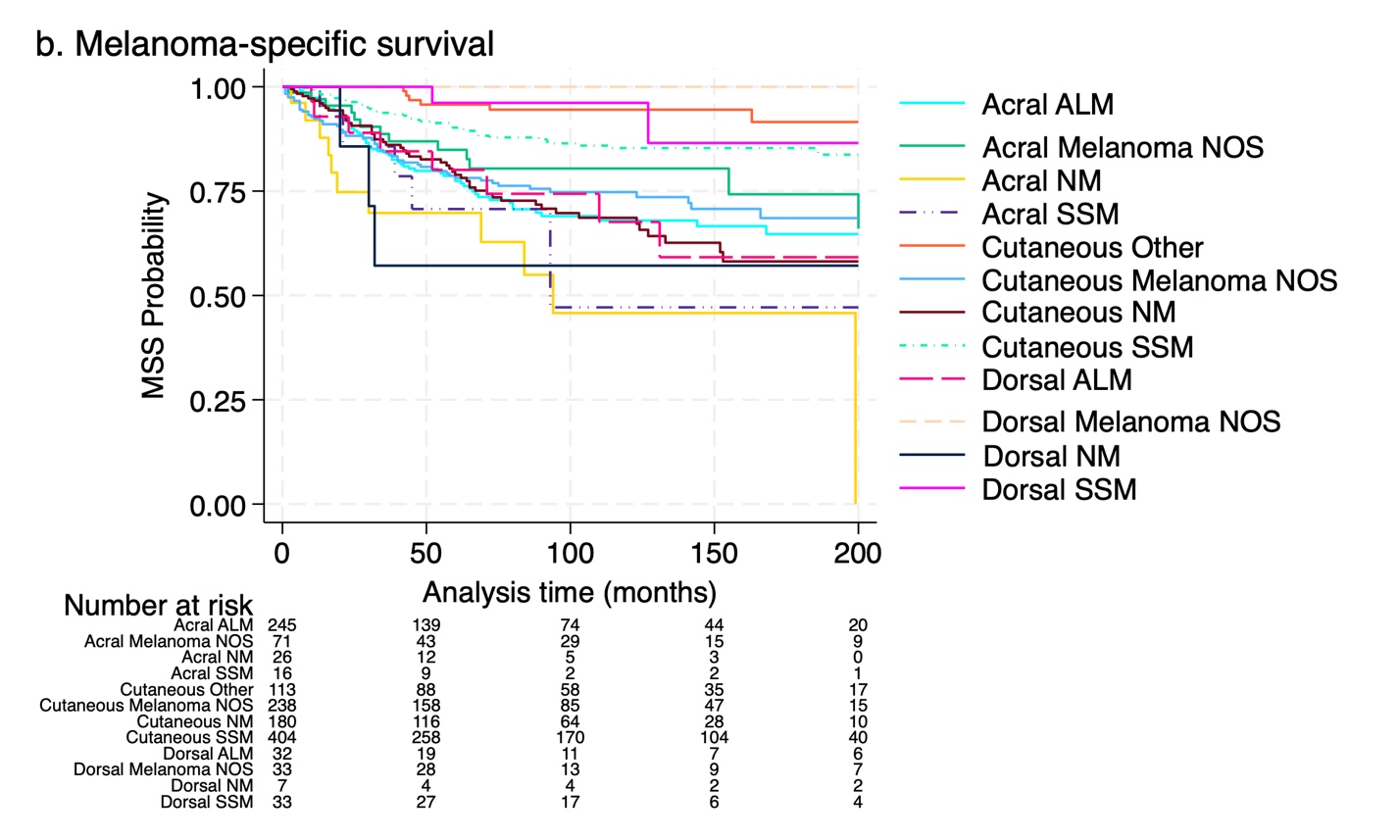

**eFigure 3c**. Kaplan-Meier curves by the interaction between location and histologic classification of melanoma for overall survival (OS). Acral location includes melanomas on palmar, plantar, and subungual surfaces. Dorsal location includes melanomas on non-subungual dorsal surfaces of the hands and feet. Cutaneous location refers to cutaneous melanomas on non-distal extremities. “Other” histology includes lentigo maligna and lentigo maligna melanoma due to low sample sizes. Acronyms: ALM=Acral lentiginous melanoma; NOS=Not otherwise specified; NM=Nodular melanoma; SSM=Superficial spreading melanoma.

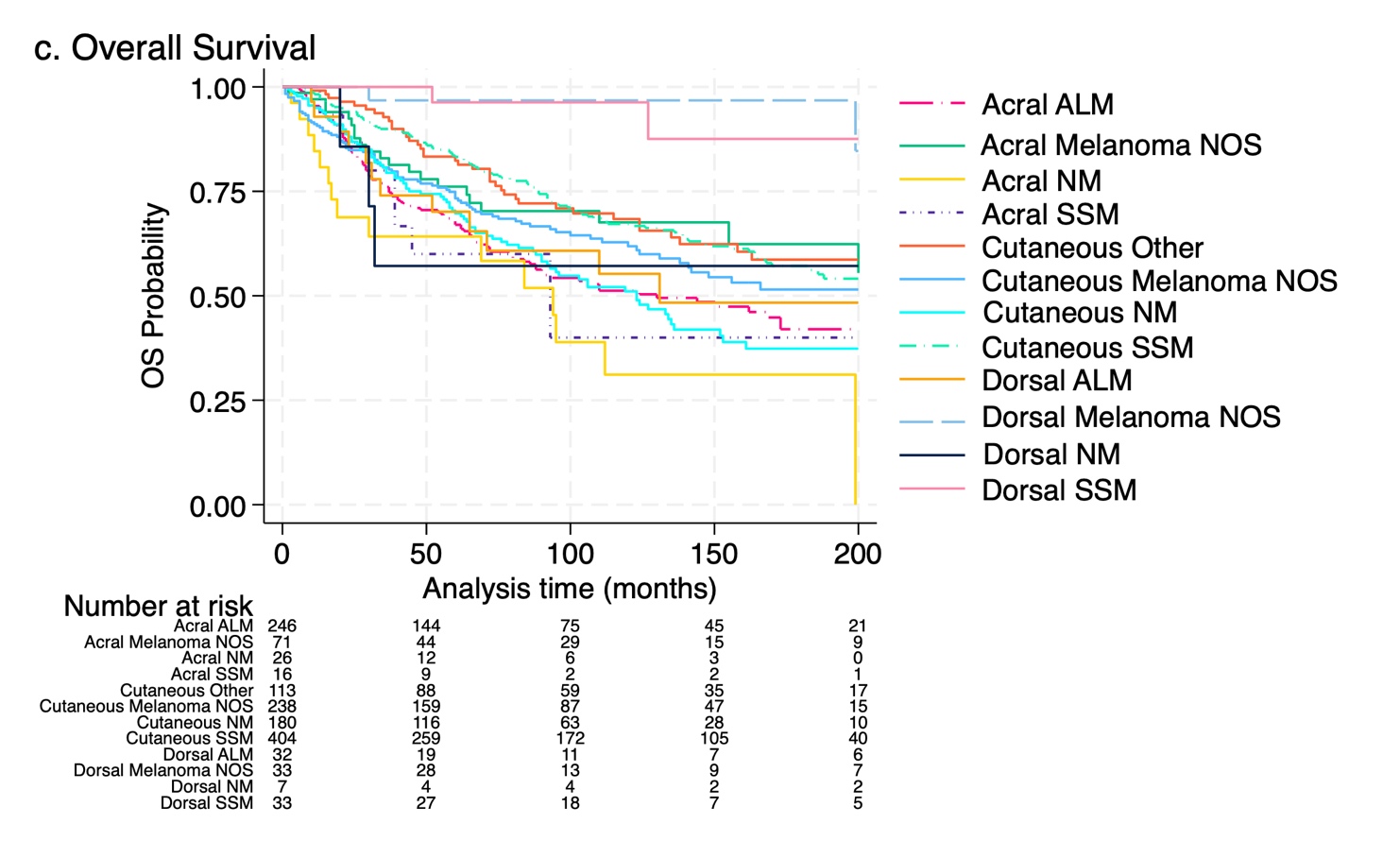
